# A Better Way: Initial Acceptability Testing of Using Artificial Intelligence Tools to Accelerate Development of Trauma Clinical Guidance

**DOI:** 10.1101/2025.08.20.25334097

**Authors:** Gabriela Zavala Wong, Shannon Rosenauer, Chelsea Church, Diana Sherifali, Megan Racey, Katheryn Grider, Ashley N. Moreno, Lacey N. LaGrone, The 2025 Design for Implementation (DFI) Authorship Group, Pamela Bixby, Stephanie Bonne, Eileen M. Bulger, James G Cain, Jennifer Chastek, Julia Roberts Coleman, Todd W Costantini, Nicholas Cozzi, Kimberly A. Davis, Rochelle A. Dicker, Warren C. Dorlac, Erik Van Eaton, Evert Eriksson, Susan Evans, Shannon Marie Foster, Jeffrey M. Goodloe, Elliott R. Haut, Molly Jarman, Alyssa Johnson, Meera Kotagal, Morgan Krause, John C. Kubasiak, Kelly Lang, Allison Barbara Leigh, Halinder S. Mangat, Debra Marie Marvel, Christopher Paul Michetti, Vicki Moran, Ashley N. Moreno, Simon JW Oczkowski, Michael A. Person, Michelle A. Price, LJ Punch, Megan Racey, Bradford L. Ray, Diane Redmond, Linda Kate Reinhart, Heather Rhodes, Bryn Rhodes, Andres M. Rubiano, Sabrina Sanchez, Babak Sarani, Erica Shelton, David A Spain, Kristan Staudenmayer, Deborah M. Stein, Julie Valenzuela, Cynthia Lizette Villarreal, Jeffrey L. Wells, Gabriela Zavala Wong, LeAnne Sitari Young

## Abstract

**Introduction:** Representatives of the trauma community have voiced a need for a new approach to developing clinical guidance. In this study, we test the initial acceptability of a proposed 12-step approach that aims to reduce the current clinical guidance timeline from more than 24 months to 24 weeks.

**Methods:** Investigators hypothesized that artificial intelligence (AI) tools could be leveraged to improve and make the process of clinical guidance development more efficient, facilitating AI initial output that could later be reviewed by subject matter experts (SMEs). Ensuring ethical standards and a collaborative design. Following the agile methodology, emphasizing continuous delivery and improvement, and the Practical, Robust Implementation and Sustainability Model (PRISM) framework, the investigators drafted a 12-step approach to clinical guidance development in 24 weeks. The process starts with the selection of a clinical topic and culminates in a bedside-ready clinical decision tree.

**Results:** The 2025 Design *for Implementation: The Future of Trauma Research & Clinical Guidance* conference participants were invited to reflect on this new 12-step approach during two breakout sessions. Participants included a broad range of trauma providers, methodologists, patient representatives, technology, and marketing experts. Their recommendations highlighted: 1) multidisciplinary involvement, 2) need for resource-stratified recommendations, and 3) user-friendly features (offline and multilingual access). On a post conference survey (n=56), 64% were confident in AI accelerating the current development process.

**Conclusions:** The current landscape of clinical guidance offers significant opportunities for improvement. Key areas for enhancement include promoting collaboration across multiple disciplines and organizations, developing recommendations that consider resource variations, and utilizing new technologies, such as AI, to expedite the development process. This is crucial because ongoing delays lead to practices lagging behind current evidence. Further research is needed to rigorously test and refine how responsible use of AI can be integrated into expediting evidence integration into clinical guidance.

**Key Messages:** *What is already known on this topic:* Current clinical guidance typically takes 1-2 years to develop. Moreover, clinical guidance may not be published until a year or more after its completion, long after some recommendations become outdated, contributing to lagged evidence-informed practice.

*What this study adds:* This study shares and tests the initial acceptability of a novel approach that aims to reduce the current clinical guidance timeline from 24 months to 24 weeks. It leverages existing artificial intelligence tools but with the critical input of subject matter experts (SMEs), ensuring ethical standards and collaborative design. SMEs shed light on critical steps and key areas that future clinical guidance needs to consider.

*How this study might affect research, practice or policy:* The current landscape of clinical guidance offers significant opportunities for improvement. Key areas for enhancement include promoting collaboration across multiple disciplines and organizations, developing recommendations that consider resource variations, and utilizing new technologies, such as artificial intelligence, to expedite the development process.

## INTRODUCTION

Clinical guidance is the cornerstone of evidence-informed practice, promoting recommendations that aid clinical decision making and reduce harmful, ineffective practices. [1] Nonetheless, there is still an existing gap between what we know and what we practice due to implementation barriers that are often ignored, including suboptimal healthcare networks, time constraints, poor applicability to real-world clinical practice, lack of equipment, and human resources. [2–3] These barriers make clinical guidance impractical and inapplicable to several settings failing to fulfill its purpose: supporting consistent quality clinical care. [4] Studies have documented this challenge across multiple regions and clinical specialties. For instance, a 2016 systematic review and meta-analysis reported a global decrease in adherence rates to venous thromboembolism prophylaxis guidance. [5] Another review on World Health Organization’s Guidelines for Essential Trauma Care, found that less than 30% of the world had implemented these guidelines to any extent. [6]

While numerous resources have been developed, their content fails to account for predictable, already identified implementation barriers. Implementation matters because as Rice et al. described in 2012, a major deviation in the standard of care for injured patients resulted in a threefold increase in 30- and 90-day mortality. [7] Similar results were found in Godier et al (2016), where clinical guidance compliance resulted in decreased mortality at 24 hours and 30 days. [8] Furthermore, a 2024 assessment of 20 trauma clinical guidance found most of them do not consider usability aspects and lack of patient-friendly resources. [9]

It is estimated that it takes an average of 17 years for 14% of evidence [10] to reach clinical practice. Furthermore, clinical guidance may not be published until a year or more after its completion, long after the publication date of most of the studies included in the clinical guidance. [11] By its publication date, newer studies or interventions may have occurred, and some recommendations will have become outdated; therefore, even if providers are following current clinical guidance, there are still knowledge gaps and a lag in evidence-informed practice. [12]

Traumatic injuries, one of the leading causes of death in the United States, are among the major health categories that suffer from the chaotic landscape of clinical guidance development. A 2024 systematic review found that most trauma clinical guidance is developed for and by high-income settings.[13] Thus, multiple organizations are currently making the effort to build more accessible, evidence-informed clinical guidance for trauma care to aid clinical decision making in austere environments where surgical workforce density and skillsets are sometimes insufficient to meet the demands safely. [14–16] External evidence is better adapted when it’s conducted in a collaborative environment that considers the variability of resources available and values local experts. [17] However, this remains a global problem, and even the most well-resourced health systems can become resource constrained due to natural disasters, civil conflicts, or technology outages.

Clinical guidance adaptation frameworks provide a step-by-step approach to implement existing clinical guidance into a specific local context. Several frameworks exist, including the Practice Guidelines Evaluation and Adaptation Cycle (PGEAC) [18], Systematic Guideline Review (SGR) [19], ADAPTE [20], The Alberta Ambassador Program (AAP) adaptation process [21], CAN-IMPLEMENT [22], SNAP-IT by GRADE (MAGIC) [23], Adapted ADAPTE [24], Appraisal of Guidelines for Research & Evaluation II (AGREE-II) [25], and GRADE-ADOLOPMENT. [26] Nonetheless, there are still gaps in the impact of clinical guidance adaptation and therefore implementation, especially in low-resource settings and low- and middle-income countries (LMICs). Harrison et al. (2013) found that it sometimes takes more than 24 months to complete the clinical guidance adaptation process. This process did not represent a faster approach than clinical guidance creation, 1-2 years. [27]

Representatives of the trauma community are voicing a need for a new approach and proposing strategies that focus on professional society collaboration, designing primary clinical research for implementation, a systematic prioritization of selected clinical topics for clinical guidance development, enhancing transparent authorship representation, planning for regular review and revision of clinical guidance, improving discoverability of clinical guidance, optimizing user experience, disseminating clinical guidance, and utilization of open access and open licenses. [28–31]

Today’s trauma care is already changing with the inclusion of artificial intelligence (AI) in healthcare. Machine learning and large language models (LLMs) are aiding healthcare practitioners with suggested therapeutics embedded in electronic health records (EHRs) [32], enhanced imaging diagnostics [33], patient monitoring [34], medical device automation [35], transcription of provider-patient encounters [36], and much more. Inspired by the needs of the trauma community and given the emerging innovations in AI, we hypothesize that AI can be leveraged, following ethical standards, to improve the current process of clinical guidance development while considering resource variability when providing evidence-informed recommendations. Thus, reducing the current timeline for clinical guidance development to 24 weeks, from topic selection to a bedside-ready clinical decision tree. This manuscript reflects the initial acceptability testing of this hypothesized approach with subject matter experts (SMEs) during the 2025 *Design for Implementation: The Future of Trauma Research & Clinical Guidance* (DFI) conference.

## METHODS

### The 12-Step Approach for Clinical Guidance Development

Based on previous findings [13,30–31], following the agile methodology [37], emphasizing continuous delivery and improvement, and the Practical, Robust Implementation and Sustainability Model (PRISM) framework [38] to scale and improve current clinical guidance development process, participants were presented with a 12-step approach (Table 1) to clinical guidance development.

**Table 1:**
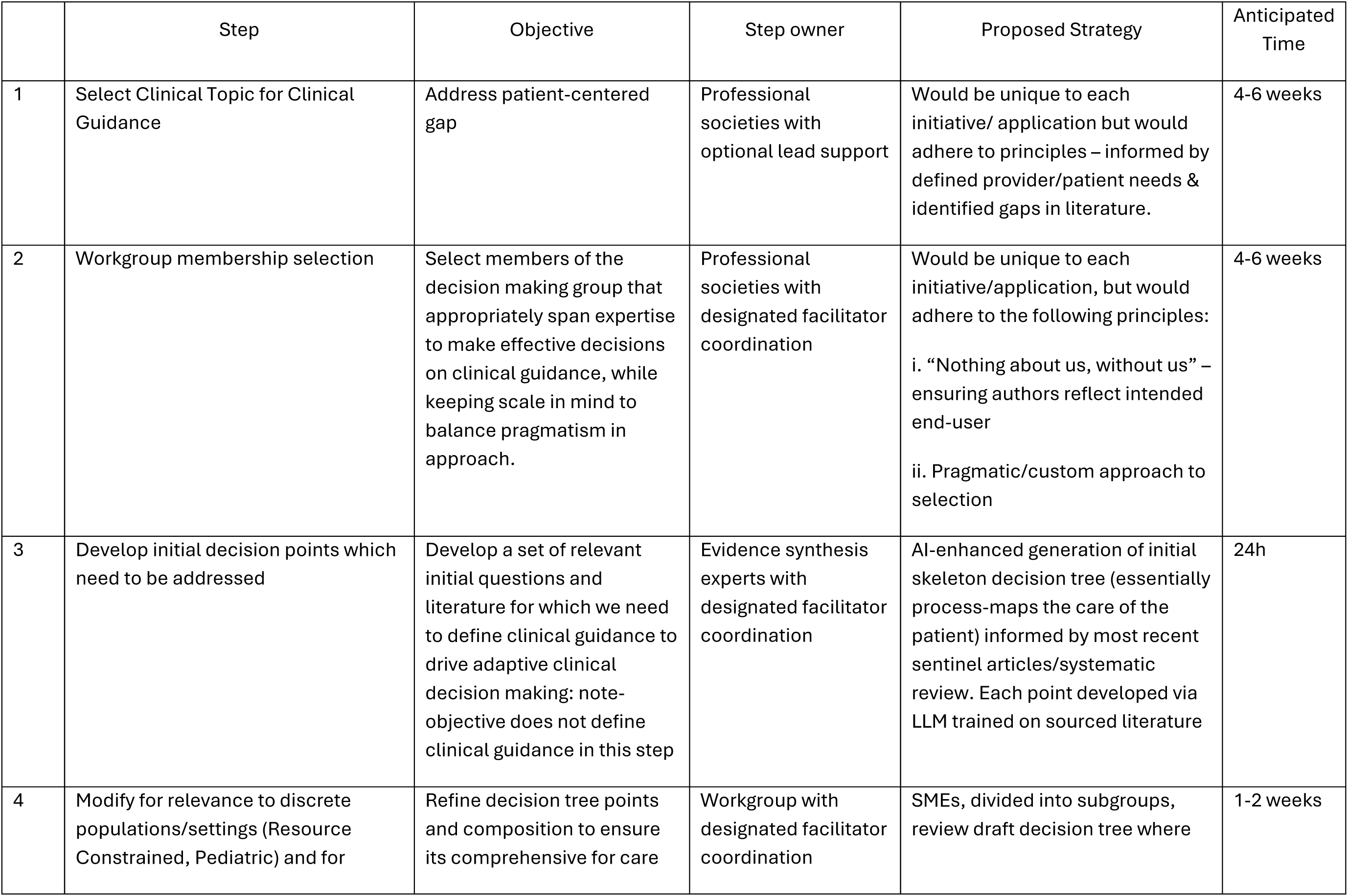
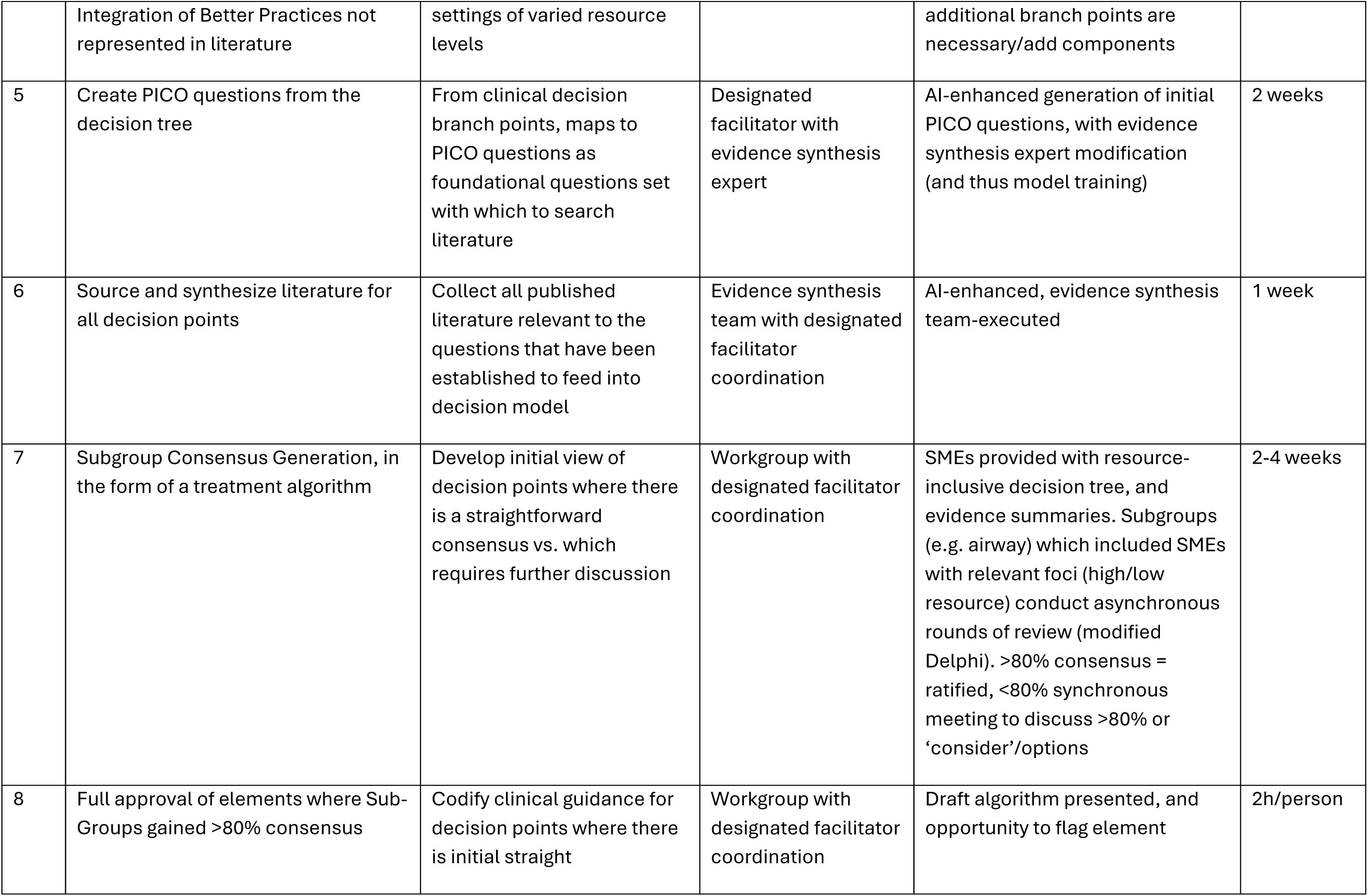
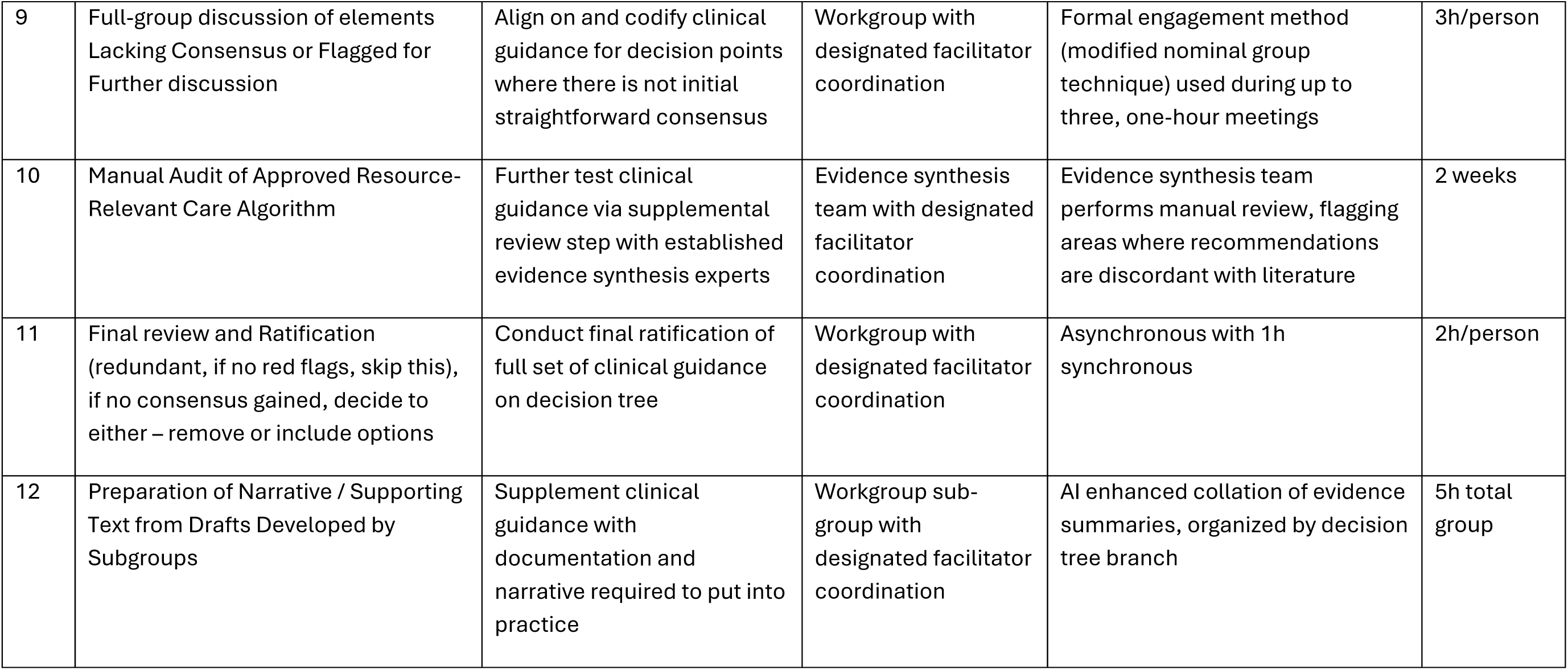
12-step approach for clinical guidance development.

|  | Step | Objective | Step owner | Proposed Strategy | Anticipated Time |
| --- | --- | --- | --- | --- | --- |
| 1 | Select Clinical Topic for Clinical Guidance | Address patient-centered gap | Professional societies with optional lead support | Would be unique to each initiative/ application but would adhere to principles – informed by defined provider/patient needs & identified gaps in literature. | 4-6 weeks |
| 2 | Workgroup membership selection | Select members of the decision making group that appropriately span expertise to make effective decisions on clinical guidance, while keeping scale in mind to balance pragmatism in approach. | Professional societies with designated facilitator coordination | Would be unique to each initiative/application, but would adhere to the following principles:<br>i. “Nothing about us, without us” – ensuring authors reflect intended end-user<br>ii. Pragmatic/custom approach to selection | 4-6 weeks |
| 3 | Develop initial decision points which need to be addressed | Develop a set of relevant initial questions and literature for which we need to define clinical guidance to drive adaptive clinical decision making: note- objective does not define clinical guidance in this step | Evidence synthesis experts with designated facilitator coordination | AI-enhanced generation of initial skeleton decision tree (essentially process-maps the care of the patient) informed by most recent sentinel articles/systematic review. Each point developed via LLM trained on sourced literature | 24h |
| 4 | Modify for relevance to discrete populations/settings (Resource Constrained, Pediatric) and for | Refine decision tree points and composition to ensure its comprehensive for care | Workgroup with designated facilitator coordination | SMEs, divided into subgroups, review draft decision tree where | 1-2 weeks |
|  | Integration of Better Practices not represented in literature | settings of varied resource levels |  | additional branch points are necessary/add components |  |
| 5 | Create PICO questions from the decision tree | From clinical decision branch points, maps to PICO questions as foundational questions set with which to search literature | Designated facilitator with evidence synthesis expert | AI-enhanced generation of initial PICO questions, with evidence synthesis expert modification (and thus model training) | 2 weeks |
| 6 | Source and synthesize literature for all decision points | Collect all published literature relevant to the questions that have been established to feed into decision model | Evidence synthesis team with designated facilitator coordination | AI-enhanced, evidence synthesis team-executed | 1 week |
| 7 | Subgroup Consensus Generation, in the form of a treatment algorithm | Develop initial view of decision points where there is a straightforward consensus vs. which requires further discussion | Workgroup with designated facilitator coordination | SMEs provided with resource-inclusive decision tree, and evidence summaries. Subgroups (e.g. airway) which included SMEs with relevant foci (high/low resource) conduct asynchronous rounds of review (modified Delphi). >80% consensus = ratified, <80% synchronous meeting to discuss >80% or 'consider'/options | 2-4 weeks |
| 8 | Full approval of elements where Sub-Groups gained >80% consensus | Codify clinical guidance for decision points where there is initial straight | Workgroup with designated facilitator coordination | Draft algorithm presented, and opportunity to flag element | 2h/person |
| 9 | Full-group discussion of elements Lacking Consensus or Flagged for Further discussion | Align on and codify clinical guidance for decision points where there is not initial straightforward consensus | Workgroup with designated facilitator coordination | Formal engagement method (modified nominal group technique) used during up to three, one-hour meetings | 3h/person |
| 10 | Manual Audit of Approved Resource-Relevant Care Algorithm | Further test clinical guidance via supplemental review step with established evidence synthesis experts | Evidence synthesis team with designated facilitator coordination | Evidence synthesis team performs manual review, flagging areas where recommendations are discordant with literature | 2 weeks |
| 11 | Final review and Ratification (redundant, if no red flags, skip this), if no consensus gained, decide to either – remove or include options | Conduct final ratification of full set of clinical guidance on decision tree | Workgroup with designated facilitator coordination | Asynchronous with 1h synchronous | 2h/person |
| 12 | Preparation of Narrative / Supporting Text from Drafts Developed by Subgroups | Supplement clinical guidance with documentation and narrative required to put into practice | Workgroup sub-group with designated facilitator coordination | AI enhanced collation of evidence summaries, organized by decision tree branch | 5h total group |

### Setting and Participants

To obtain feedback and gather reflections from SMEs, we engaged key partners (i.e., trauma providers, professional society leadership, methodological experts, patient representatives, technology experts, and marketing experts) during the second annual DFI conference that occurred February 19-20, 2025, in Chicago, Illinois, USA. Convenience snowball sampling was employed for recruiting in-person participants. This study was reviewed by the Colorado Multiple Institutional Review Board, CB F490; COMIRB No: 24-1608 and COMIRB #: 22-0626 and determined exempt from institutional review board review. Seventy people participated in-person, and up to 55 virtual participants attended via Zoom.

### Conference Structure and Activities

Conference attendees participated in two breakout sessions where each one of the 12 steps was explained, and they were invited to provide low-fidelity process feedback through a set of questions relating to each step (see Supplemental Item 1). The objective of these sessions was to reflect on the approach to a combined synchronous/asynchronous, AI-accelerated, SME-informed, evidence synthesis expert and ethicists-regulated process to the development of resource-relevant clinical guidance with a beside-side ready decision tree. Participants provided their responses either verbally or via an anonymous polling platform (Slido; https://www.slido.com/). In a semi-structured focus group format, follow-up questions were conducted when participants raised concerns and shared improvement strategies to improve portions of this proposed 12-step process.

After these two breakout groups concluded, an additional subgroup of nine SMEs was selected to pilot test this new strategy. The nine participants within the SME subgroup were selected based on their research, experience with, and high involvement in, clinical guidance development. Efforts were taken to ensure this group contained adequate representation of patient advocates, physicians, nurses, and methodological experts – simulating the recommended approach for real-world development of clinical guidance.

Participants were then presented with a draft algorithm which had been developed using out-of-the-box AI tools. For piloting, the volume resuscitation portion of a damage control resuscitation algorithm was selected. [39] This was chosen for recency of publication, and representativeness of a national trauma collaboration. The full-text references from this algorithm were then loaded into Chat Generative Pre-Trained Transformer (GPT)-4 Plus and a query written to prompt Chat-GPT to draft a clinical algorithm. The output was reviewed and query revised until the algorithm drafted presented content of basic clinical utility and accuracy. In sequence, prompts requested Chat-GPT to summarize references, create bullet-point recommendations, and convert them into a clinical decision tree.

This AI-generated volume resuscitation clinical decision tree was presented in an interactive platform (Miro; www.miro.com/app) to allow for live-editing and integration of user-feedback. SMEs were first asked to review asynchronously, using mobile survey tooling to either agree with each node of the presented algorithm, or suggest modification. Then, facilitators took the group through the results of the asynchronous polling and the narrative text of the suggested modifications.

### Post-Conference Survey

Finally, an anonymous post-conference survey was administered to all participants via Microsoft Forms (Microsoft Corporation, 2024) to capture final thoughts on this new 12-step approach to clinical guidance creation and the future of clinical guidance development through Likert-scale questions.

### Data Analysis

SME breakout sessions feedback was captured via an anonymous polling platform, Slido (https://www.slido.com/). Through inductive coding, SMEs’ reflections were grouped into thematic categories. These included: Patient Advocacy, Workgroup Selection, Project Management, Workload Organization, Dissemination and Implementation Science, Funding Opportunities, Information and Technology, Diverse Language, AI Ethical Standards, Expert Opinion, Aligned Incentives, Accountability Resource Variability, Consensus Methodology, Quantity Cut-Off of Consensus and Mutual Understanding of Consensus Disagreement and National Incentives.

Participants’ quotes were included for better representation of each category. SME subgroup voting and comments were also gathered via Slido. Their voting results were expressed as percentages, to facilitate interpretation and identification of consensus areas among respondents. When SMEs disagreed with one of the recommendations included in the clinical decision tree, they were encouraged to submit their modification request via Slido as a comment. These underwent inductive coding and were grouped into the following thematic categories: Patient Advocacy, Accountability for Resource Variability, Need for Detailed Recommendations, Accountability for Different Populations and Format. Post-conference survey results were presented using descriptive statistics.

## RESULTS

### Perspectives of the 12-step Approach

Seventy in-person, and up to 55 virtual participants included a broad range of SMEs, including trauma providers, professional societies leadership, methodological experts, patient representatives, and technology and marketing experts. When the 12-step approach was presented during two breakout sessions, SMEs provided their input, and not all steps triggered observations or comments (Table 2). SMEs expressed the desire to include more patients and patient advocates; assured diverse representation across healthcare providers and practice settings (e.g., low-, middle- and high-resource settings; rural and urban) and shared concerns with lack of engagement and time constraints. They also highlighted the need for more user-friendly features like offline content availability, mobile app-based medical information, information in more languages, and open-access information. They agreed about the importance of reviewing AI-augmented steps and following implementation and dissemination strategies to achieve sustainability.

**Table 2.**
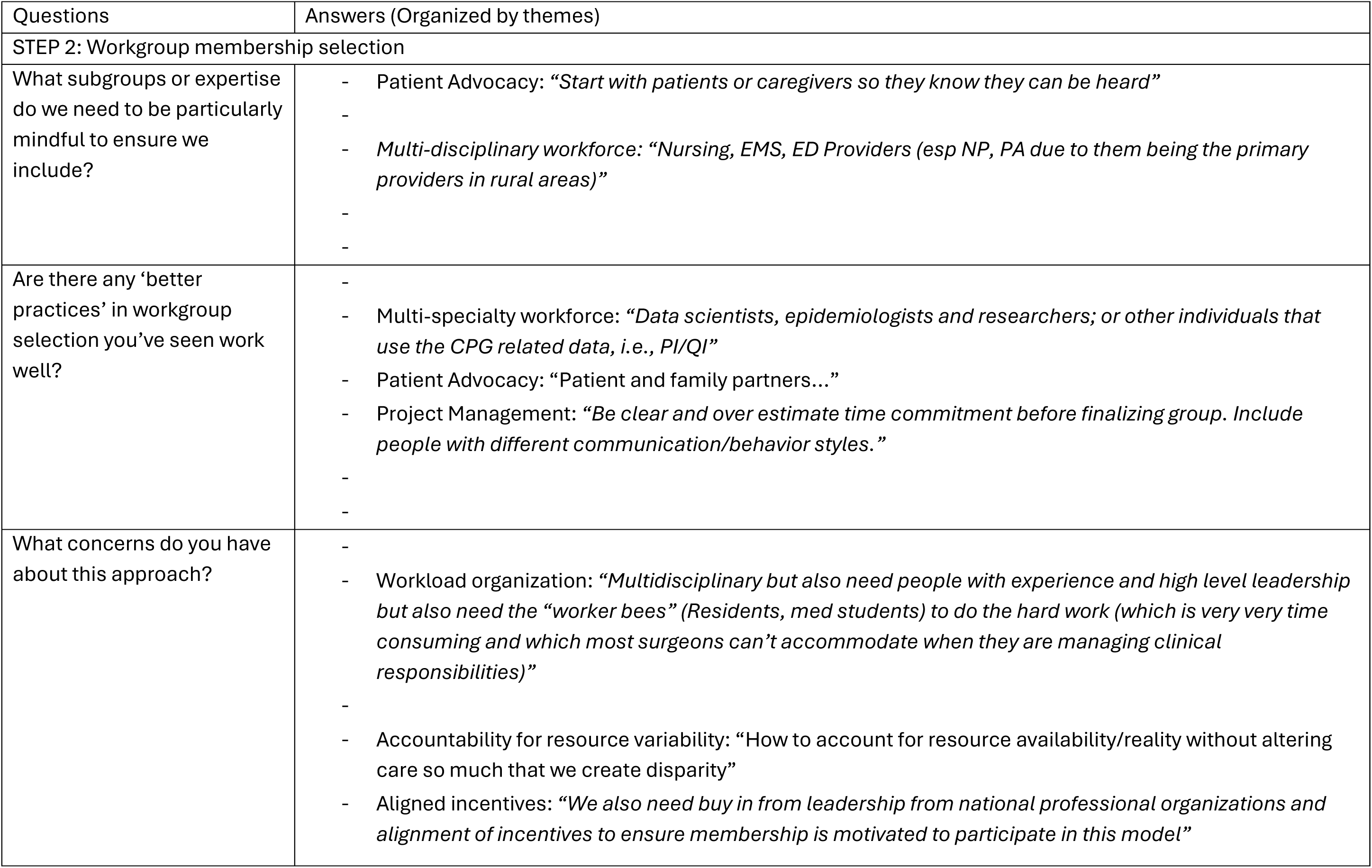
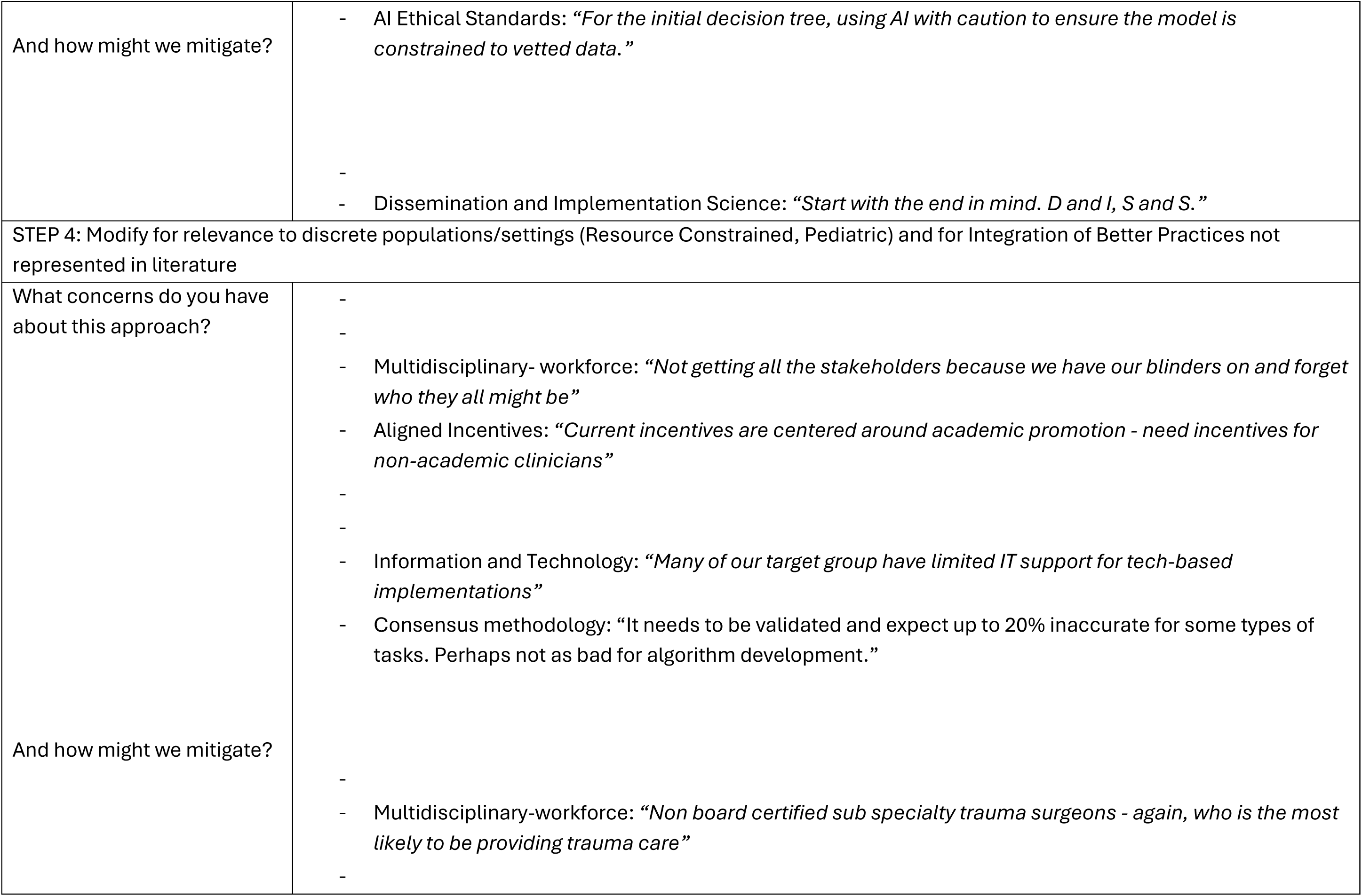
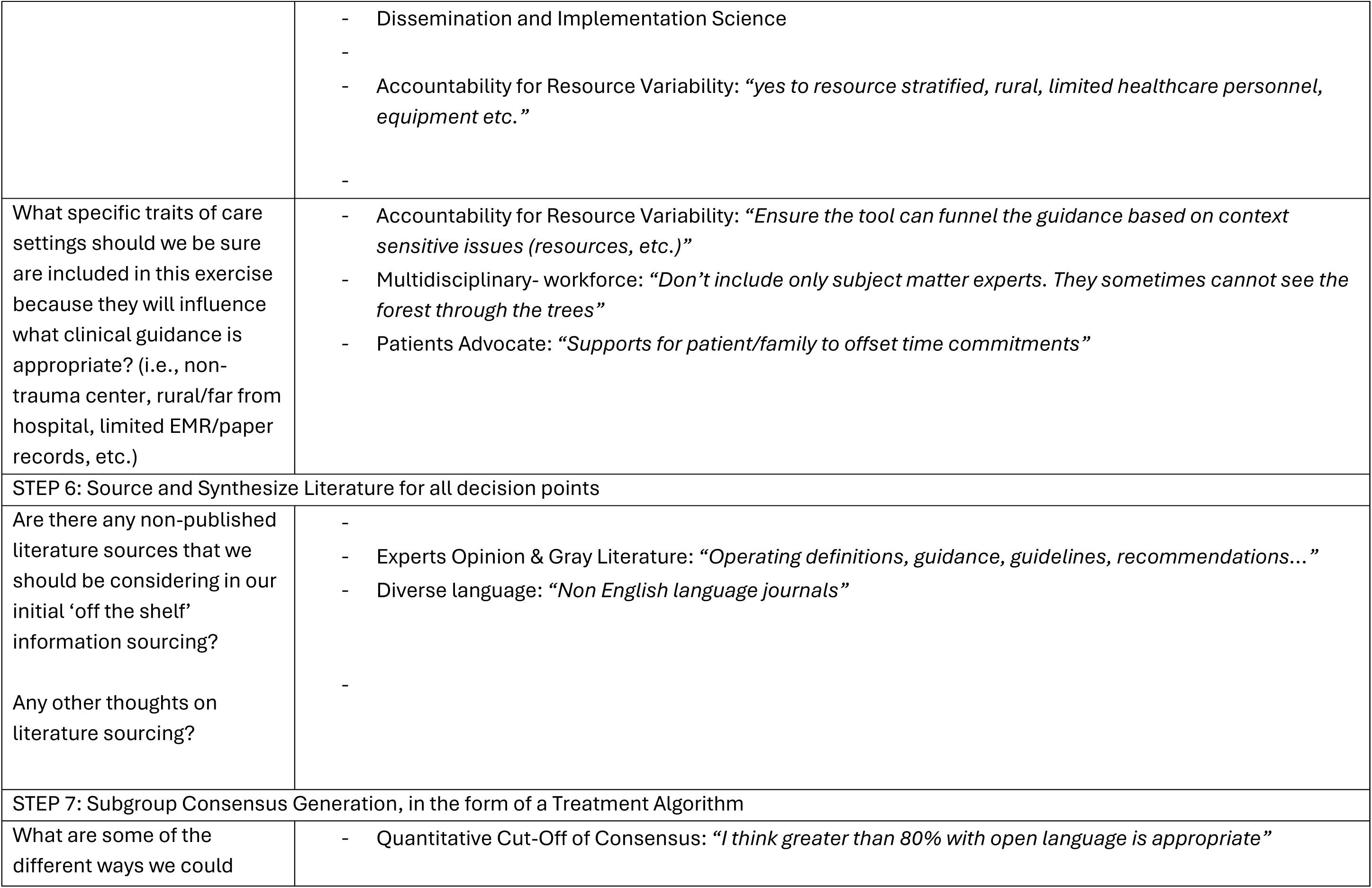
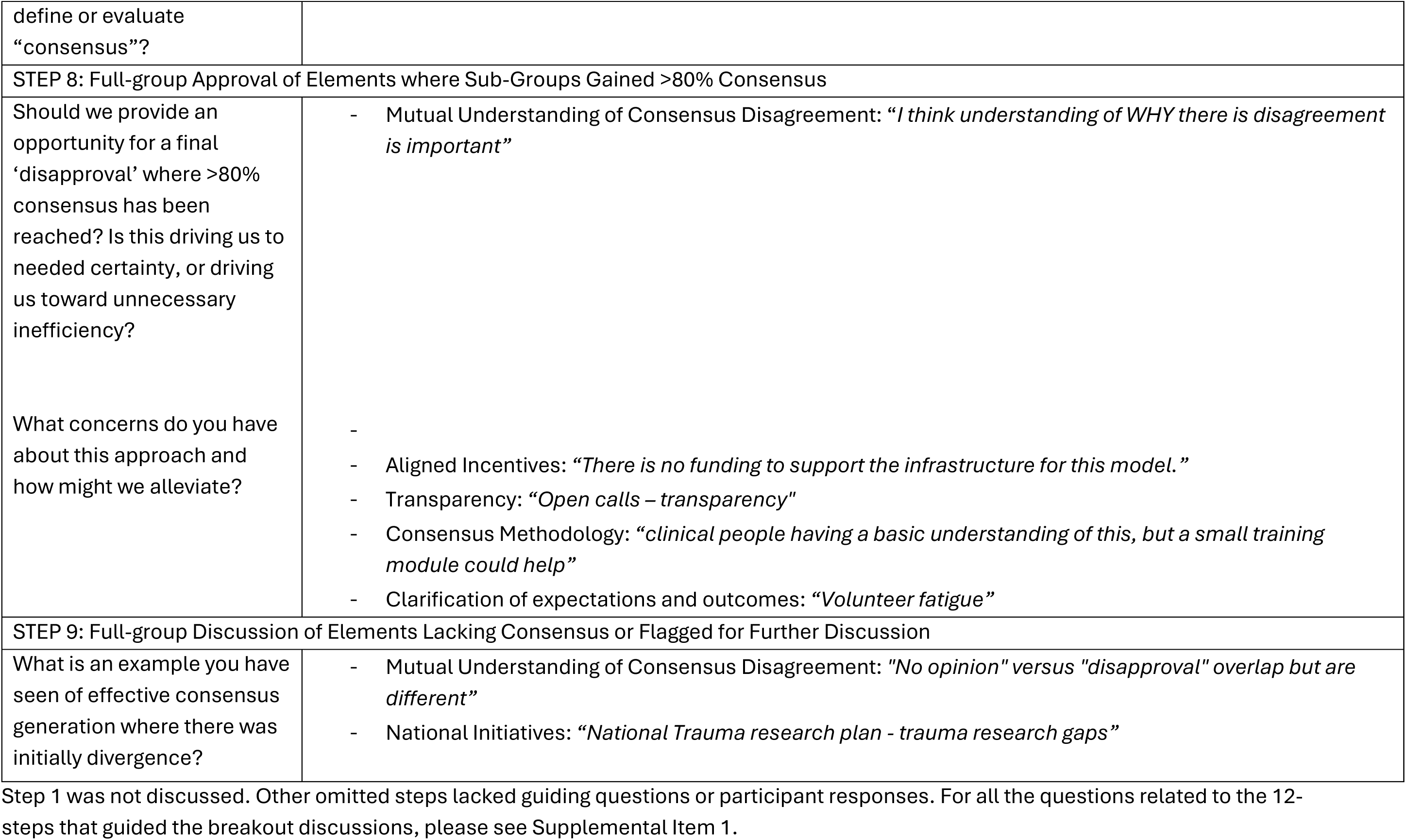
SMEs Breakout Sessions Reflecting on 12-Step Approach.

During the pilot test of a preliminary algorithm (Figure 1) for volume resuscitation created using AI augmentation with the smaller, nine person subgroup of SMEs (one patient advocate, three methodological experts and five trauma providers), participant feedback highlighted once again the need to include additional decision tree nodes (pathways) that reflected resource variability, equipment, and trained personnel, suggested more specific recommendations and more information about treatment options (e.g., range, age, when, how and what to monitor, inclusion criteria, comorbidities and lab values), the inclusion of additional population thresholds, and advised on formatting and avoiding repetitive or redundant recommendations and connectors (see Table 3 and Figure 2). Most recommendations had less than 60% of agreement. Blood pressure parameters were the only recommendations that had 100% agreement; the recommendations that triggered less agreement (22%) were found to be redundant or had missing information.

**Figure 1.**
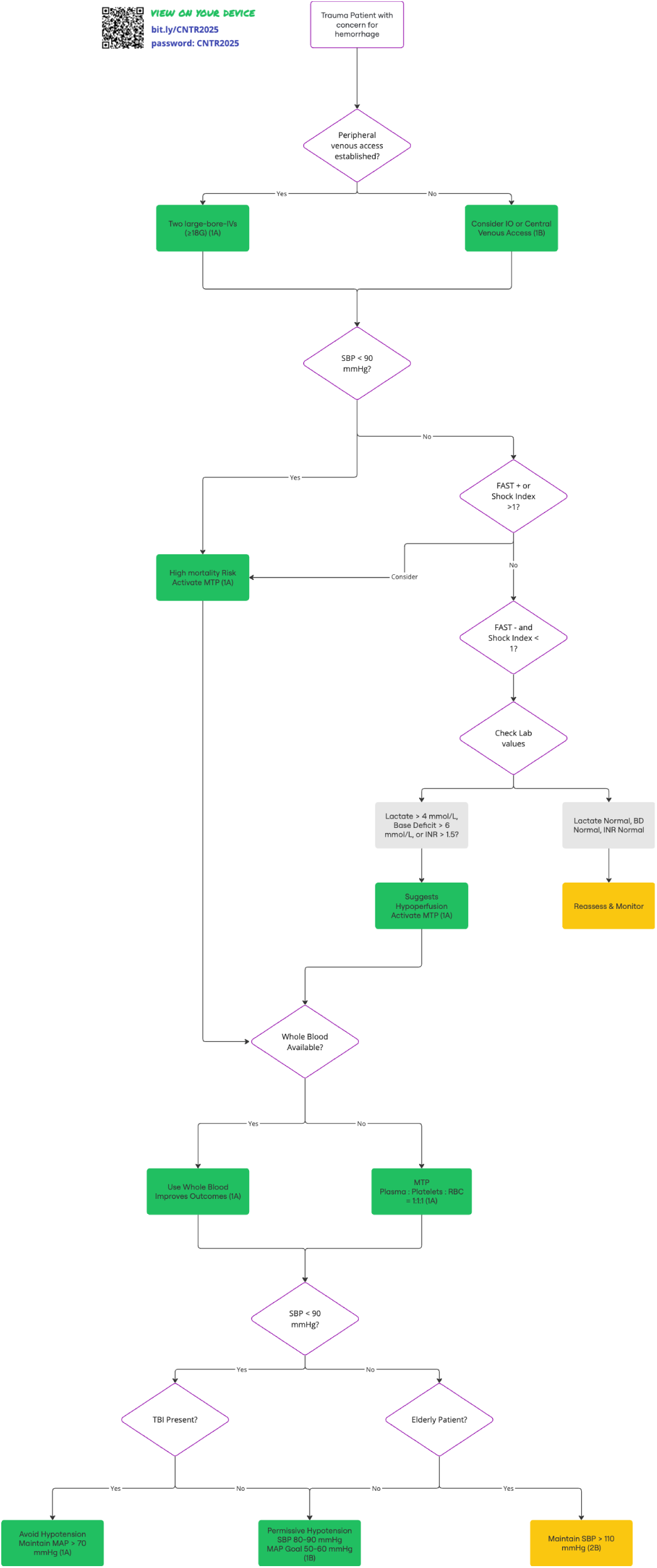
AI-augmented volume resuscitation algorithm

**Figure 2.**
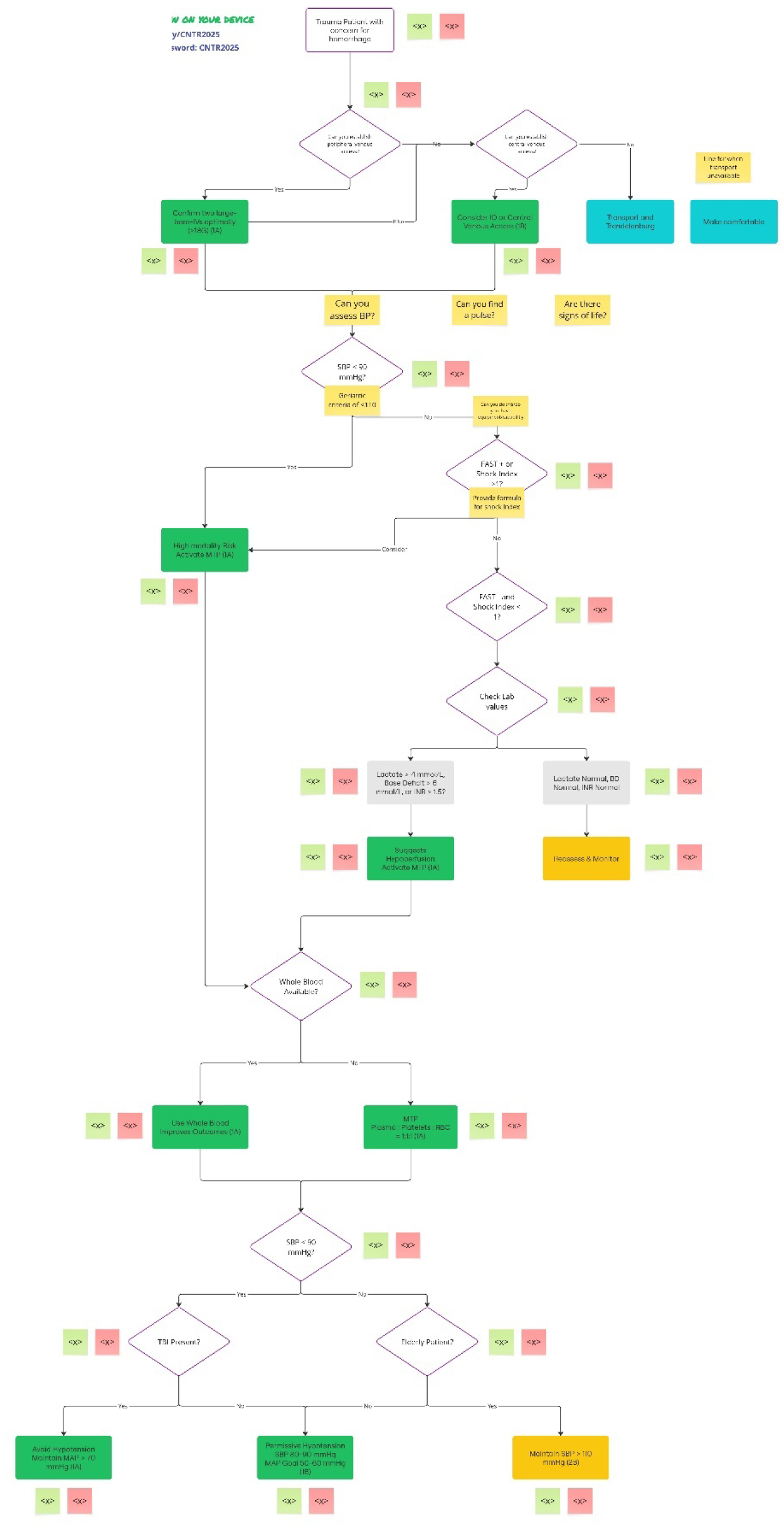
AI-augmented volume resuscitation algorithm incorporating SME’s feedback

**Table 3:**
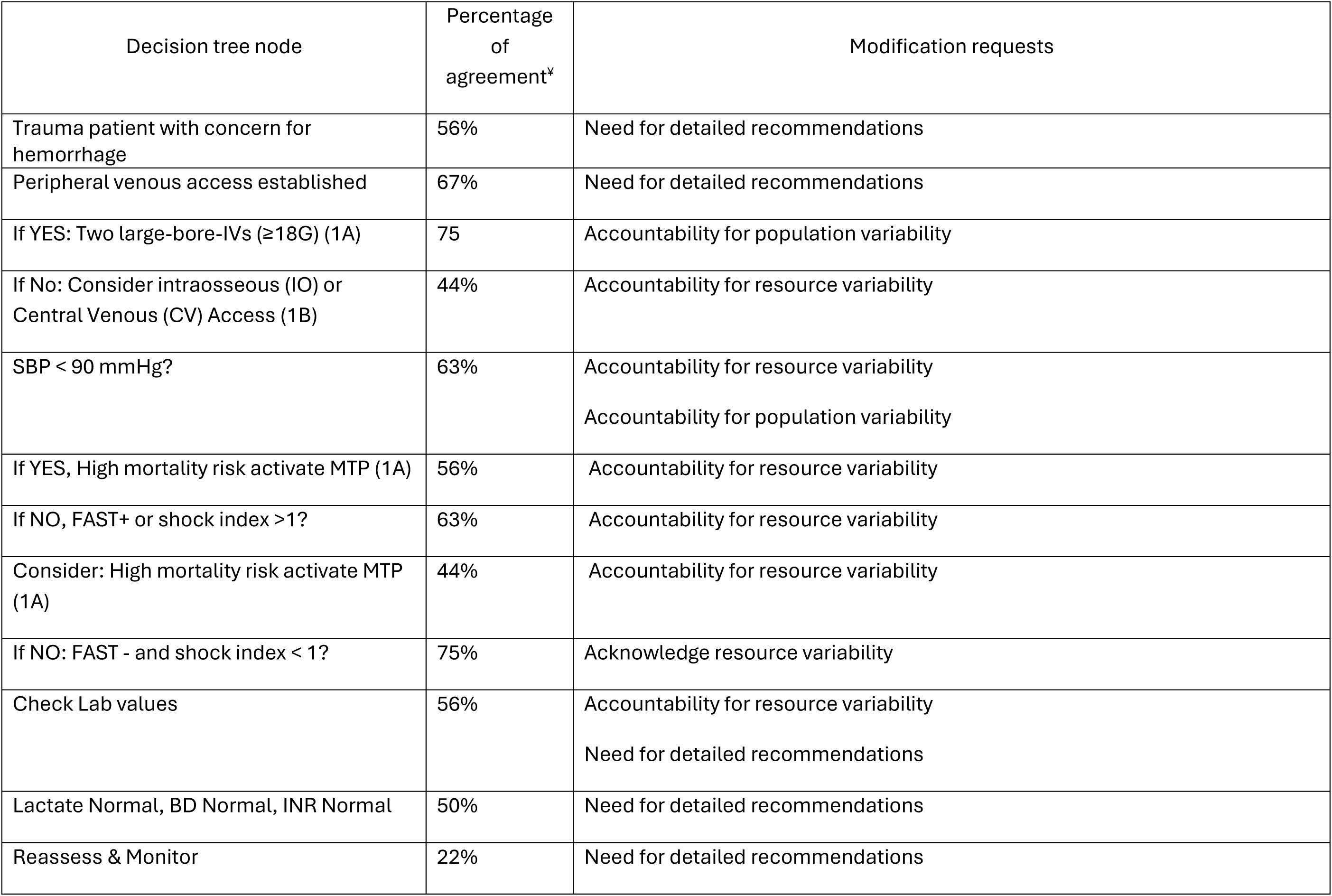
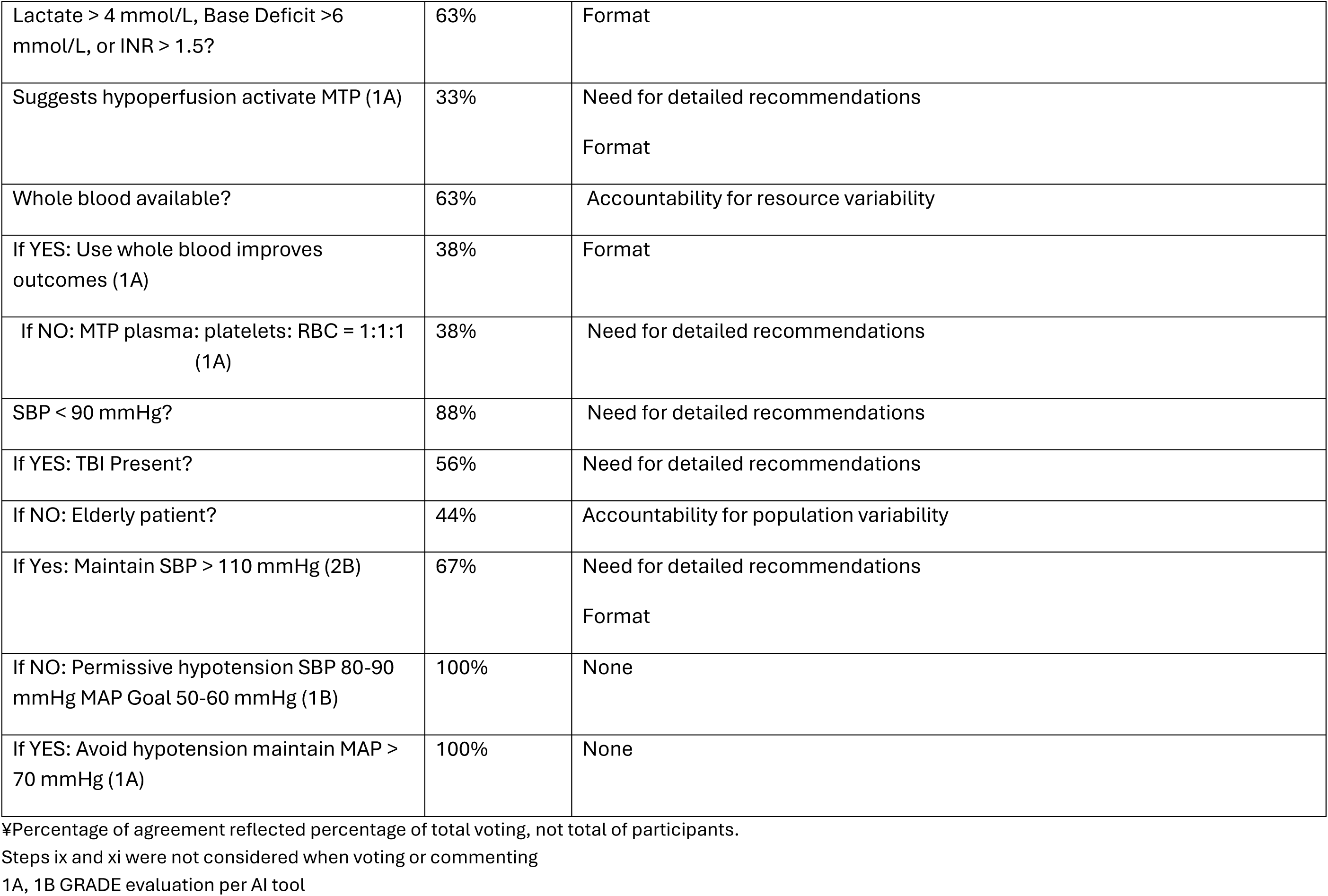
SME’s pilot test AI-augmented algorithm.

| Decision tree node | Percentage of agreement* | Modification requests |
| --- | --- | --- |
| Trauma patient with concern for hemorrhage | 56% | Need for detailed recommendations |
| Peripheral venous access established | 67% | Need for detailed recommendations |
| If YES: Two large-bore-IVs ($\geq 18G$) (1A) | 75 | Accountability for population variability |
| If No: Consider intraosseous (IO) or Central Venous (CV) Access (1B) | 44% | Accountability for resource variability |
| SBP < 90 mmHg? | 63% | Accountability for resource variability<br>Accountability for population variability |
| If YES, High mortality risk activate MTP (1A) | 56% | Accountability for resource variability |
| If NO, FAST+ or shock index >1? | 63% | Accountability for resource variability |
| Consider: High mortality risk activate MTP (1A) | 44% | Accountability for resource variability |
| If NO: FAST - and shock index < 1? | 75% | Acknowledge resource variability |
| Check Lab values | 56% | Accountability for resource variability<br>Need for detailed recommendations |
| Lactate Normal, BD Normal, INR Normal | 50% | Need for detailed recommendations |
| Reassess & Monitor | 22% | Need for detailed recommendations |
| Lactate > 4 mmol/L, Base Deficit >6 mmol/L, or INR > 1.5? | 63% | Format |
| Suggests hypoperfusion activate MTP (1A) | 33% | Need for detailed recommendations<br>Format |
| Whole blood available? | 63% | Accountability for resource variability |
| If YES: Use whole blood improves outcomes (1A) | 38% | Format |
| If NO: MTP plasma: platelets: RBC = 1:1:1 (1A) | 38% | Need for detailed recommendations |
| SBP < 90 mmHg? | 88% | Need for detailed recommendations |
| If YES: TBI Present? | 56% | Need for detailed recommendations |
| If NO: Elderly patient? | 44% | Accountability for population variability |
| If Yes: Maintain SBP > 110 mmHg (2B) | 67% | Need for detailed recommendations<br>Format |
| If NO: Permissive hypotension SBP 80-90 mmHg MAP Goal 50-60 mmHg (1B) | 100% | None |
| If YES: Avoid hypotension maintain MAP > 70 mmHg (1A) | 100% | None |

¥Percentage of agreement reflected percentage of total voting, not total of participants.
Steps ix and xi were not considered when voting or commenting
1A, 1B GRADE evaluation per AI tool

### Post-Conference Survey

In a final reflection, 56 participants provided their thoughts on the future of clinical guidance development via the post-conference survey. Seventy-nine percent (n=44) of respondents believed that the current status quo of clinical decision making leaves room for improvement. Despite the algorithm being a beta, very preliminary prototype of what the future of clinical guidance could be, 64% of respondents were confident that AI can be useful synthesizing complex clinical guidance to increase translation of evidence into practice. Finally, more than 70% of participants rated user-friendliness, graphics, ability to personalize settings, relevance, and ethical considerations as important factors for a bedside-ready clinical guidance tool.

## DISCUSSION

The research team proposed a 12-step approach to improve the current status of clinical guidance development, leveraging LLMs and existing AI to reduce the current burden SMEs experience with today’s approach. Nonetheless, the value of SMEs’ input as irreplaceable was highlighted, and having AI-augmented steps requires setting standards and regulatory reviews to ensure quality throughout the development process. After reviewing this new approach with SMEs during the conference, the importance of ensuring adequate representation (e.g. rural, urban, high-, medium- and low-resource settings), multidisciplinary groups of healthcare providers and scientists, patient advocates, muti-system/multi-organizational coordination, and a standardized framework to follow was essential. SMEs’ concerns revolved around lack of time, lack of funding, sustainability, project coordination and management, adaptability to reflect resource variability, and implementation and dissemination of the developed products. When a subgroup of SMEs tested a very preliminary AI-augmented clinical decision tree on volume resuscitation, their review became more granular in terms of content, wording, and formatting; however, the critical need to include resource variability within the clinical decision support was again highlighted.

As previously discussed, the current clinical guidance development is far from ideal, and today’s clinical practice has a more than 10-year gap [8]; additionally, the time invested in clinical guidance development is between 1-2 years. [11,40] Prioritizing the improvement of this process is imperative as clinical decision making benefits from consistent, quality care through clinical guidance. Further, the current timeline fails to scale when a pandemic, socio-political conflict, or natural disaster demands clinical decision support that considers resource variability and the unknown nature of a new disease. Some organizations have started to collaborate and join in efforts for a more practical way to develop clinical guidance. The British Medical Journal (BMJ), Australian Living Evidence Collaboration, World Health Organization (WHO), and Stroke Foundation have partnered with MAGIC Evidence Ecosystem Foundation to develop rapid, trustworthy recommendations to improve the delivery of quality care. [41] An example is the BMJ Rapid Recommendation on Low Intensity Pulsed Ultrasound (LIPUS) for fracture healing. [42]

Advances in evidence synthesis and technology are promising for the future of clinical guidance. Khraisha et al. (2024) evaluated the performance of GPT-4, one of the biggest LLMs on the market, on title/abstract screening, full-text review, and data extraction across various literature types and languages. Results were variable in screening and data extraction; however, with screening full-text literature, its performance reached “human-like” levels. [43] Our proposed approach builds on LLMs, accelerating the process in a structured manner that ensures regulatory oversight and SME governance to deliver quality and accountability.

Finally, SMEs, in line with current literature, voiced that clinical guidance development requires multidisciplinary partnerships to guarantee the creation of relevant recommendations that are rigorous and high quality. A cross-disciplinary approach harnesses a variety of contexts (diverse populations, cultures, healthcare systems) to create more applicable, adaptable, and implementable clinical guidance. [44–45] Guidelines International Network (GIN) exemplifies the collaboration efforts of 93 organizations and 89 individuals representing 46 countries that aim to set international standards for clinical guidance development. [45]

### Limitations

Most of the participants came from urban, academic, and high-resource backgrounds. Although virtual participation was intentionally open and not restricted to such environments, representation from rural, low-resources, community-based settings was unbalanced; and, therefore, could be a potential bias when capturing SMEs perspectives. To mitigate this, when piloting the AI-augmented algorithm, the research team selected a more balanced representation of SMEs, taking into consideration urban, rural, academic, and non-academic experts. It is also recognized that the sample size was small overall and results reflect low-fidelity feedback. In addition, integration and use of AI in clinical guidance development remains in early stages, and requires further refinement, validation, and standardization. While the proposed approach aims to incorporate resource variability, the research team recognizes that it is not fully limited to trained personnel or infrastructure constraints but encompasses healthcare norms and system-level practices that could differ significantly across countries. These factors were not explicitly addressed in this initial model.

## CONCLUSION

The current landscape of clinical guidance offers significant opportunities for improvement. Key areas for enhancement include promoting collaboration across multiple disciplines and organizations, developing recommendations that consider variations in resources, and utilizing innovative technologies, such as AI, to expedite the development process. This is crucial because ongoing delays lead to lagged evidence-informed practice. New approaches, including the one herein proposed, to creating clinical guidance merit further research to assess and refine models rigorously. Such efforts would enhance consistency, relevance, implementability, and timeliness of the clinical guidance, ultimately improving care across various settings.

## SUPPLEMENTAL ITEMS

Supplemental Item 1. Breakout Session Slides

## Data Availability

A limited, deidentified subset of data produced in the present study are available upon reasonable request to the authors.

## References

1. Committee on Standards for Developing Trustworthy Clinical Practice Guidelines, Board on Health Care Services, Institute of Medicine. Clinical Practice Guidelines We Can Trust [Internet]. Graham R, Mancher M, Wolman DM, Greenfield S, Steinberg E, editors. Washington, D.C.: National Academies Press; 2011

2. Guerra-Farfan E, Garcia-Sanchez Y, Jornet-Gibert M, et al. Clinical practice guidelines: The good, the bad, and the ugly. Injury. 2023 May;54:S26–9.

3. Wang T, Tan JY (Benjamin), Liu XL, Zhao I. Barriers and enablers to implementing clinical practice guidelines in primary care: an overview of systematic reviews. BMJ Open. 2023 Jan;13(1):e062158.

4. Zhou P, Chen L, Wu Z, et al. The barriers and facilitators for the implementation of clinical practice guidelines in healthcare: an umbrella review of qualitative and quantitative literature. Journal of Clinical Epidemiology. 2023 Oct;162:169–81.

5. Farfan M, Bautista M, Bonilla G, Rojas J, Llinás A, Navas J. Worldwide adherence to ACCP guidelines for thromboprophylaxis after major orthopedic surgery: A systematic review of the literature and meta-analysis. Thrombosis Research. 2016 May;141:163–70.

6. LaGrone L, Riggle K, Joshipura M, Quansah R, Reynolds T, Sherr K, et al. Uptake of the World Health Organization’s trauma care guidelines: a systematic review. Bull World Health Organ. 2016 Aug 1;94(8):585–598C.

7. Rice TW, Morris S, Tortella BJ, et al. Deviations from evidence-based clinical management guidelines increase mortality in critically injured trauma patients*: Critical Care Medicine. 2012 Mar;40(3):778–86.

8. Godier A, Bacus M, Kipnis E, et al. Compliance with evidence-based clinical management guidelines in bleeding trauma patients. British Journal of Anaesthesia. 2016 Nov;117(5):592–600.

9. Wilson DJ, Zavala Wong G, Tignanelli C, et al. TRAUMA: making trauma clinical guidance more implementable. Trauma Surg Acute Care Open 2025;0:e001610.

10. Beauchemin M, Cohn E, Shelton RC. Implementation of Clinical Practice Guidelines in the Health Care Setting: A Concept Analysis. Advances in Nursing Science. 2019 Oct;42(4):307–24.

11. Morris ZS, Wooding S, Grant J. The answer is 17 years, what is the question: understanding time lags in translational research. J R Soc Med. 2011 Dec;104(12):510–20.

12. Shekelle P, Woolf S, Grimshaw JM, et al. Developing clinical practice guidelines: reviewing, reporting, and publishing guidelines; updating guidelines; and the emerging issues of enhancing guideline implementability and accounting for comorbid conditions in guideline development. Implementation Sci. 2012 Dec;7(1):62.

13. Zavala Wong G, Panshin MS, Samsamshariat T, et al. Systematic review of global trauma clinical guidance: an assessment of accessibility, relevance and quality. Trauma Surg Acute Care Open 2025;0:e001624.

14. Rubiano AM, Vera DS, Montenegro JH, et al. Recommendations of the Colombian Consensus Committee for the Management of Traumatic Brain Injury in Prehospital, Emergency Department, Surgery, and Intensive Care (Beyond One Option for Treatment of Traumatic Brain Injury: A Stratified Protocol [BOOTStraP]). J Neurosci Rural Pract. 2020 Jan;11(01):007–22.

15. Di Saverio S, Birindelli A, Kelly MD, et al. WSES Jerusalem guidelines for diagnosis and treatment of acute appendicitis. World J Emerg Surg. 2016 Dec;11(1):34.

16. Mock C, International Society of Surgery, editors. Guidelines for essential trauma care. Geneva: World Health Organization; 2004. 93 p. (Services).

17. Sørensen JB, Housseine N, Maaløe N, et al. Scaling up Locally Adapted Clinical Practice Guidelines for Improving Childbirth Care in Tanzania: A Protocol for Programme Theory and Qualitative Methods of the PartoMa Scale-up Study. Global Health Action. 2022 Dec 31;15(1):2034136.

18. Graham ID, Harrison MB. Evaluation and adaptation of clinical practice guidelines. Evid Based Nurs. 2005 July;8(3):68–72.

19. Muth C, Gensichen J, Beyer M, et al. The Systematic Guideline Review: Method, rationale, and test on chronic heart failure. BMC Health Serv Res. 2009 Dec;9(1):74.

20. The ADAPTE Collaboration. The ADAPTE Process: Resource Toolkit for Guideline Adaptation. 2009. Available from: http://www.g-i-n.net.

21. Harstall C, Taenzer P, Angus DK, et al. Creating a multidisciplinary low back pain guideline: anatomy of a guideline adaptation process. Evaluation Clinical Practice. 2011 Aug;17(4):693–704.

22. Harrison MB, van den Hoek J, the Canadian Guideline Adaptation Study Group. CAN-IMPLEMENT© guideline adaptation and implementation planning resource. Kingston, Ontario, Canada: Queen’s University School of Nursing and Canadian Partnership Against Cancer; 2012.

23. Kristiansen A, Brandt L, Agoritsas T, et al. Adaptation of Trustworthy Guidelines Developed Using the GRADE Methodology. Chest. 2014 Sept;146(3):727–34.

24. Amer YS, Elzalabany MM, Omar TI, et al. The ‘Adapted ADAPTE’: an approach to improve utilization of the ADAPTE guideline adaptation resource toolkit in the A lexandria Center for Evidence-Based C linical Practice Guidelines. Evaluation Clinical Practice. 2015 Dec;21(6):1095–106.

25. Brouwers MC, Kho ME, Browman GP, Burgers JS, Cluzeau F, Feder G, et al. AGREE II: advancing guideline development, reporting and evaluation in health care. Canadian Medical Association Journal. 2010 Dec 14;182(18):E839–42.

26. Schünemann HJ, Wiercioch W, Brozek J, Etxeandia-Ikobaltzeta I, Mustafa RA, Manja V, et al. GRADE Evidence to Decision (EtD) frameworks for adoption, adaptation, and de novo development of trustworthy recommendations: GRADE-ADOLOPMENT. Journal of Clinical Epidemiology. 2017 Jan;81:101–10.

27. Harrison MB, Graham ID, Van Den Hoek J, et al. Guideline adaptation and implementation planning: a prospective observational study. Implementation Sci. 2013 Dec;8(1):49.

28. LaGrone LN, Stein DM, Wilson DJ, et al. Equitable and effective clinical guidance development and dissemination: trauma aims to lead the way. Trauma Surg Acute Care Open. 2024 Dec;9(1):e001338.

29. Wilson DJ, Gellings JA, Zavala G, et al. Conference proceedings for the 2024 design for implementation: the future of trauma research and clinical guidance conference series. Trauma Surg Acute Care Open 2024;0:e001583.

30. Steiner A, Person MA, Bowe D, et al. Understanding the rural injury providers’ experiences with trauma clinical guidance – a qualitative case series. Trauma Surg Acute Care Open 2025;0:e001598. doi:10.1136/tsaco-2024-001598

31. Guido JM, Krause M, Steiner A, et al. Trauma community clinical guidance needs: a mixed-methods iterative consensus building study. Trauma Surg Acute Care Open 2025;0:e001592. doi:10.1136/tsaco-2024-001592

32. Yang R, Tan TF, Lu W, et al. Large language models in health care: Development, applications, and challenges. Health Care Science. 2023 Aug;2(4):255–63.

33. Hosny A, Parmar C, Quackenbush J, et al. Artificial intelligence in radiology. Nat Rev Cancer. 2018 Aug;18(8):500–10.

34. Dubey A, Tiwari A. Artificial intelligence and remote patient monitoring in US healthcare market: a literature review. Journal of Market Access & Health Policy. 2023 Dec 31;11(1):2205618.

35. U.S Food & Drug Administration [Internet]. 2025. Artificial Intelligence-Enabled Medical Devices.

36. Tierney AA, Gayre G, Hoberman B, et al. Ambient Artificial Intelligence Scribes to Alleviate the Burden of Clinical Documentation. NEJM Catalyst [Internet]. 2024 Feb 21;5(3).

37. Association for Project Management [Internet]. What is agile project management?

38. Feldstein AC, Glasgow RE. A Practical, Robust Implementation and Sustainability Model (PRISM) for Integrating Research Findings into Practice. The Joint Commission Journal on Quality and Patient Safety. 2008 Apr;34(4):228–43.

39. LaGrone LN, Stein D, Cribari C, et al. American Association for the Surgery of Trauma/American College of Surgeons Committee on Trauma: Clinical protocol for damage-control resuscitation for the adult trauma patient. J Trauma Acute Care Surg. 2024 Mar;96(3):510–20.

40. IDSA Standards and Practice Guidelines Committee. Handbook for Clinical Practice Guidelines Development. Infectious Diseases Society of America; 2021. 74 p.

41. MAGIC Evidence Ecosystem Foundation [Internet]. 2025. Available from: https://www.magicevidence.org/

42. Poolman RW, Agoritsas T, Siemieniuk RAC, Harris IA, Schipper IB, Mollon B, et al. Low intensity pulsed ultrasound (LIPUS) for bone healing: a clinical practice guideline. BMJ. 2017 Feb 21;j576.

43. Khraisha Q, Put S, Kappenberg J, Warraitch A, Hadfield K. Can large language models replace humans in systematic reviews? Evaluating GPT-4’s efficacy in screening and extracting data from peer-reviewed and grey literature in multiple languages. Research Synthesis Methods. 2024 July;15(4):616–26.

44. Korylchuk N, Pelykh V, Nemyrovych Y, et al. Challenges and Benefits of a Multidisciplinary Approach to Treatment in Clinical Medicine. J Pioneer Med Sci. 2024 June 30;13(3):1–9.

45. Qaseem A, Forland F, Macbeth F, et al. Guidelines International Network: Toward International Standards for Clinical Practice Guidelines. Ann Intern Med. 2012 Apr 3;156(7):525–31.

